# Data-augmented machine learning redefines the effective concentration of eculizumab in complement blood disorders

**DOI:** 10.64898/2025.12.13.25342192

**Authors:** Lucía Alfonso-González, Francisco J. Fernández, Manuel Praga, Kaare Mikkelsen, M. Cristina Vega

**Author notes:** **Correspondence:** Francisco J. Fernández, M. Cristina Vega. Equally contributing senior authors (Francisco J. Fernández, M. Cristina Vega).

## Abstract

Eculizumab, a humanized monoclonal antibody targeting the complement lytic pathway protein C5, has demonstrated high efficacy in the treatment of paroxysmal nocturnal hemoglobinuria, atypical hemolytic uremic syndrome, generalized myasthenia gravis, and neuromyelitis optica spectrum disorder. However, recent reports have highlighted patients who exhibit a lack of treatment response, necessitating an increase in the recommended dose or a reduction in the dosing interval. In this study, we employed machine-learning predictive models to identify the optimal blood concentration of eculizumab to inhibit the complement lytic pathway. Additionally, we examined the impact of data augmentation through the generation of artificial data on the predictive performance of these models. In conclusion, our machine learning model predicts that the target blood concentration of eculizumab should be increased to a minimum of 152-162 μg mL–1 (up from 50-100 μg mL–1) to achieve a more complete inhibition of the complement system’s lytic pathway.

---

Eculizumab (Soliris®) is a humanized monoclonal antibody that binds complement protein C5, blocking its cleavage and preventing activation of the lytic pathway of the complement system^1^. The drug has regulatory approval from both the US FDA and the EMA for the treatment of the rare complement-mediated disorders paroxysmal nocturnal hemoglobinuria (PNH; ORPHA: 67038), atypical hemolytic uremic syndrome (aHUS; ORPHA: 213653), generalized myasthenia gravis (gMG; ORPHA: 512), and neuromyelitis optica spectrum disorder (NMOSD; ORPHA: 71291)^2^. Across these indications, eculizumab has demonstrated efficacy in reducing disease activity and improving patient outcomes^2^. The most prominent safety concern is the increased susceptibility to invasive infection by *Neisseria meningitidis*, necessitating vaccination and ongoing vigilance^3^. Additional long-term adverse effects, such as hepatotoxicity, are increasingly reported and warrant systematic evaluation^4,5^.

The optimal serum concentration of eculizumab required for sustained inhibition of the complement lytic pathway remains uncertain. The widely cited therapeutic range of 50-100 µg/ml is not derived from robust pharmacokinetic-pharmacodynamic studies and has therefore been questioned^1,6^. In clinical practice, both under- and overexposure have been observed. Subtherapeutic trough levels are associated with reduced efficacy and recurrence of disease manifestations^7^, whereas the clinical consequences of overexposure are less well defined^8^. Nonetheless, cases of drug accumulation and excessively high trough levels, particularly in children, have been documented^8,9^. This uncertainty is compounded by the extraordinary cost of therapy, estimated at US $700,000 per patient annually. These factors underscore the need for precise strategies to determine effective dosing, minimizing the risk of underdosing while avoiding unnecessary overexposure.

Advances in data-driven technologies are transforming healthcare by enabling machine learning (ML) models that improve diagnostic accuracy, reduce costs, and support personalized treatment strategies. Yet these benefits are often limited by access to large and diverse datasets, especially in rare diseases, where clinical evidence is inherently sparse. Synthetic data generation provides a solution: artificially created datasets that reproduce the statistical properties of real patient data while expanding sample size and diversity. Used responsibly, synthetic augmentation can enhance model robustness and reduce bias while safeguarding patient privacy. This approach is particularly important for complement-mediated rare disorders, where the scarcity of trial participants and the heterogeneity of disease symptoms make traditional data collection inadequate for reliable pharmacological modeling.

We have reassessed the serum concentration of eculizumab needed to inhibit complement activation in rare hematological disorders. Using publicly available clinical data from small multicenter studies across four rare indications (**Supplementary Table S1**), we trained supervised ML models to predict the likelihood of complete C5 inhibition based on drug concentration. Acknowledging the data scarcity inherent in rare diseases, we employed synthetic data augmentation to improve model robustness and generalizability. Our study adheres to the TRIPOD-AI checklist (**Supplementary Table S2**).

Clinical data on serum levels of eculizumab (total drug) and free (unbound) C5 from PNH, aHUS, gMG, and NMOSD were used as inputs (390 independent observations) for training, validation and testing of various supervised ML classification algorithms (**Fig. 1a**). Observations were labelled as Inhibition = True when free C5 concentrations were ≤0.5 μg/ml, and Inhibition = False when >0.5 μg/ml. This threshold has been previously validated as an indicator of complete complement inhibition^13^. To estimate the concentration of eculizumab required for sustained inhibition, classifiers were queried with input values ranging from 10 to 1700 μg/ml, and the lowest concentration yielding ≥80% predicted probability of inhibition was recorded. Classifier performance was evaluated using standard metrics of predictive accuracy (**Table 1**). Among the models tested, K-Nearest Neighbors (KNN) achieved the highest overall performance, with an average accuracy of 0.90 and an ROC AUC of 0.89.

**Table 1.** Comparison of the performance of machine learning models trained and tested on real data. Results are provided considering 10-fold cross-validation.

| ML model | Complement inhibition = F |  |  | Complement inhibition = T |  |  |
| --- | --- | --- | --- | --- | --- | --- |
|  | Precision | Recall | F1 sc. | Precision | Recall | F1 sc. |
| K-Nearest Neighbors | **0.95** | 0.93 | **0.94** | 0.77 | **0.83** | **0.80** |
| Extreme Gradient Boosting | 0.94 | 0.92 | 0.93 | 0.74 | 0.80 | 0.77 |
| Logistic regression | 0.94 | 0.91 | 0.92 | 0.71 | 0.78 | 0.75 |
| Gradient Boosting | 0.93 | 0.91 | 0.92 | 0.72 | 0.76 | 0.74 |
| Random Forest | 0.93 | 0.91 | 0.92 | 0.71 | 0.75 | 0.73 |
| Support Vector Machines | 0.88 | 0.93 | 0.91 | 0.70 | 0.57 | 0.63 |
| Gaussian Naïve Bayes | 0.85 | **0.97** | 0.90 | **0.79** | 0.39 | 0.52 |

**Figure 1.**
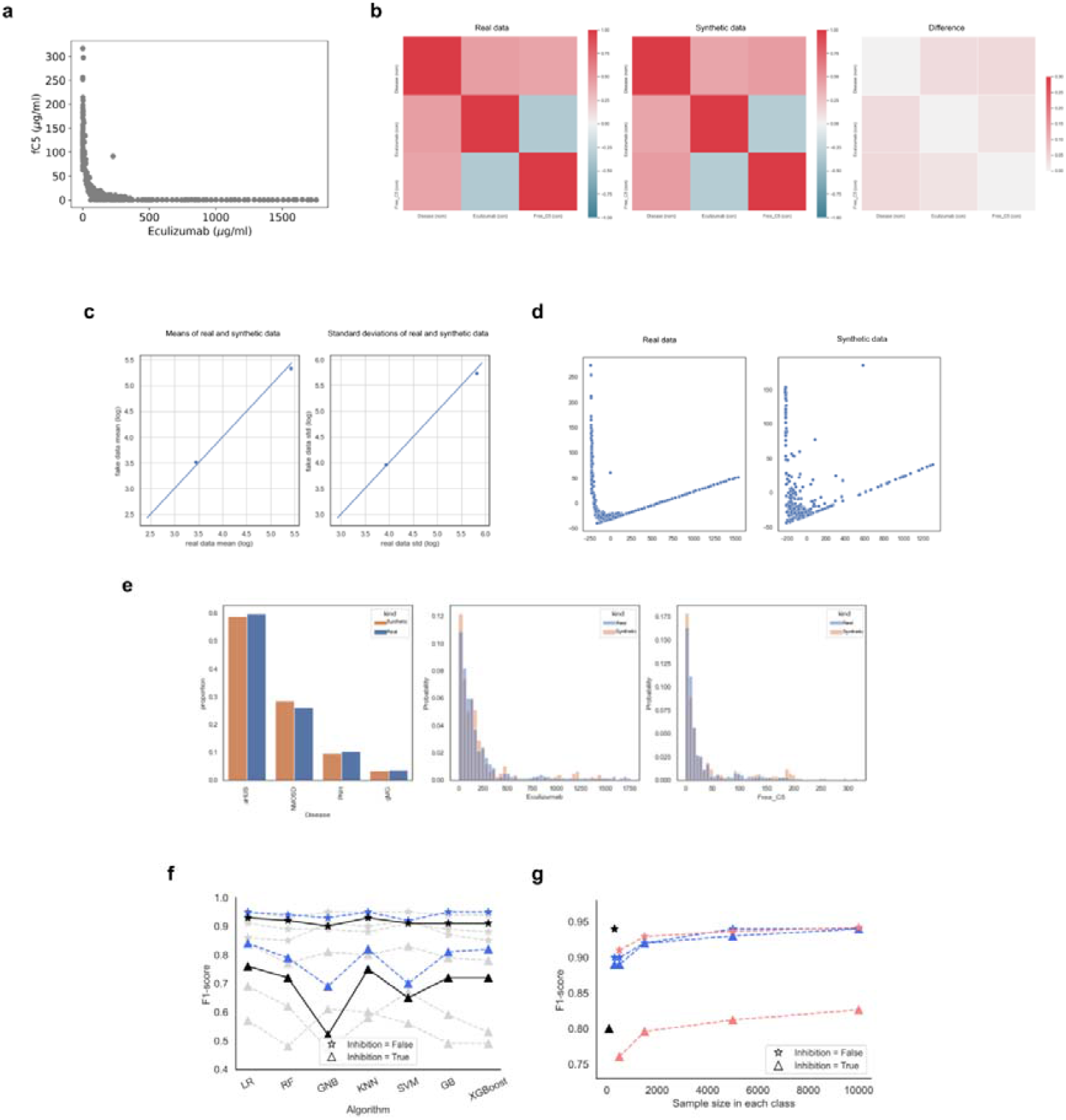
Predictive model of eculizumab maintenance concentration and evaluation of classifier performance with and without synthetic data. (a) Data on blood concentration of eculizumab and free (unbound) C5 (fC5). (b-e) Evaluation of MOSTLY-AI synthetic dataset based on its similarity to real data using statistical techniques: (b) feature correlation, (c) absolute logarithmic mean and standard deviation of numeric data, (d) first components of Principal Component Analysis, and(e) distribution per feature. Analysis made using the TableEvaluator Python library (version 1.5.0). (f) Evaluation of the MOSTLY-AI dataset compared to real data as a function of data utility. The MOSTLY-AI-generated dataset (blue) was compared with the synthetic dataset generated using other techniques (gray) and the real dataset (black), based on the F1-score of classifiers constructed using the same algorithms - Logistic Regression (LR), Random Forest (RF), Gaussian Naïve Bayes (GNB), K-Nearest Neighbors (KNN), Support Vector Machine (SVM), Gradient Boosting (GB) and Extreme Gradient Boosting (XGBoost). (g) Model performance evolution in real and synthetic data combinations. Evolution of the F1-score of models trained and tested on synthetic data in combination with real data (blue); and trained on synthetic data and tested on real data (coral), in comparison with the F1-score obtained in models trained and tested on real data (black).

The predictive ability of ML models depends on data quantity and quality. To address the limitations of sample size, we created synthetic data using multiple methods and assessed the augmented datasets for both statistical accuracy and downstream predictive performance (**Fig. 1b-f**). Synthetic data generated with MOSTLY AI most closely matched real distributions and was chosen for further analyses (**Fig. 1g**). Combining real and synthetic datasets improved classifier accuracy across all algorithms (**Supplementary Table S3**). To examine the impact of synthetic augmentation alone, we also trained models only on synthetic data and tested them on real data (**Fig. 1g**). As the dataset size grew, the prediction of the Inhibited class improved, reaching an accuracy and ROC AUC of 0.91 and 0.92, respectively, at *N* = 10,000.

Using this augmented framework, the best-performing classifiers predicted that complete complement blockade is achieved with serum eculizumab concentrations above 162 µg/ml (KNN) or 152 µg/ml (Random Forest) (**Supplementary Table S3**). To provide a robust and transparent estimate, we applied a consensus modeling strategy consistent with OECD QSAR validation principles and FDA/EMA recommendations for model-informed drug development. We therefore report a consensus threshold of 157 µg/ml, with an inter-model range of 152-162 µg/ml, as the effective maintenance concentration required for reliable C5 inhibition. Since dosing regimens for rare diseases are sensitive to miscalibration, data-augmented ML-based predictions should help mitigate the risk of clinical underdosing in current eculizumab maintenance regimens.

Our findings align with previous reports indicating higher thresholds than the commonly cited 50-100 µg/ml. For example, Reiss et al.^14^ linked concentrations above 124 µg/ml with complete blockade, while Latour et al.^15^ observed no hemolysis episodes in patients above 150 µg/ml. Clinical trial data in aHUS (and gMG) also report mean serum concentrations above 116-150 µg/ml during effective treatment^16^. Substantial inter-patient variability further complicates dosing. Cases of aHUS patients and up to 16% of PNH patients have been reported with subtherapeutic trough levels under standard regimens^15,17–19^, whereas others exhibit concentrations >500 µg/ml despite guideline-based dosing^8,20,21^. This variability underscores the importance of monitoring trough levels and adjusting both dose and interval to avoid treatment failure or overexposure. Together, these results support revising the traditional target range upwards to ensure consistent therapeutic efficacy.

Limitations of our study stem from the inherent “small-n” barrier that characterizes rare diseases and constrains the robustness of meta-analyses. Methodologically, using supervised ML introduces the risk of overfitting, which can be through cross-validation and consensus modeling. Synthetic data augmentation, while deliberately applied to overcome sample scarcity, also introduces assumptions: artificially generated datasets may not fully represent the biological and clinical variability of real patients, especially in underrepresented groups. Our analysis focused on serum eculizumab and free C5 concentrations as primary predictors; other potentially relevant biomarkers (e.g., sC5b9, or patient-specific genetic factors) were not included and could impact the true dose-response relationship. Moreover, the available data and the design of this model do not allow extrapolation of the target concentration achieved with a given dosing regimen. These limitations emphasize the need for prospective validation with larger, more diverse patient populations, ideally incorporating mechanistic complement models, such as C-model, and real-world pharmacokinetic and pharmacodynamic data, which may further enhance interpretability and clinical applicability.

ML approaches, combined with carefully validated synthetic data, offer a powerful way to fill knowledge gaps in rare-disease treatments. In this context, simulated data are crucial for training ML on rare diseases due to the limited availability of clinical data, while also protecting patients’ privacy. By expanding small training datasets while maintaining their statistical structure, this strategy allows for more accurate, clinically relevant predictions and supports model-based drug development in situations where traditional data collection is difficult. Additionally, this approach provides a scalable template for improving other therapeutics with limited trial data.

## Data Availability

All data produced in the present work are contained in the manuscript.

## Acknowledgments

MCV acknowledges the CSIC Network of Rare Diseases (RER-CSIC). LAG acknowledges support from the PhD program in Medicinal Chemistry at the Universidad Complutense de Madrid (UCM) (Spain) and a research visiting fellowship at Aarhus University (Denmark).

## Authorship

MCV and FJF conceived and designed the study. LAG, FJF, and MCV drafted and revised the work for intellectual content and context. KM analyzed the machine learning algorithms and data. MP contributed to the study’s conception and design and revised the manuscript to ensure the clinical relevance of the results. All authors contributed critically to the manuscript. MCV and FJF obtained the resources, administered, and supervised the project, and are responsible for the published work.

## Data availability

All data analyzed in this study are included in this article and its supplementary information files.

## Code availability

The custom code developed for this study may be shared with qualified researchers upon reasonable request to the corresponding author.

