## Supplementary Information for "Data-augmented machine learning redefines the effective concentration of eculizumab in complement blood disorders"

[1]Centro de Investigaciones Biológicas Margarita Salas (CIB-CSIC), Consejo Superior de Investigaciones Científicas, 28040 Madrid, Spain.

[2]Abvance Biotech SL, Pharmacokinetics, Pharmacodynamics & Drug Metabolism (PPDM), 28003 Madrid, Spain.

[3]Department of Medicine, Complutense University, 28040 Madrid, Spain.

[4]Aarhus University, Department of Electrical and Computer Engineering, 8000 Aarhus, Denmark.

*** Correspondence:**
Francisco J. Fernández

M. Cristina Vega


[#]Equally contributing senior authors

**Table of contents**

Supplementary Methods (pp. 2-3)

Supplementary Table S1. Eculizumab data (pp. 4-12).

Supplementary Table S3. TRIPOD-AI checklist (pp. 13-14)

Supplementary Table S2. Performance and predictions of ML classifiers (pp. 15-16)

References (p. 17)

### Supplementary Methods

*Data collection*

Data on blood concentrations of eculizumab and related free (unbound) C5 levels were compiled from clinical studies examining the pharmacokinetics, pharmacodynamics, efficacy, and safety of the drug (NCT00844545, NCT00838513, NCT01997229, NCT01892345, NCT02946463, NCT03056040). The final dataset included 390 independent observations (**Supplementary Table S1**). Since complete datasets were not consistently available in tabular form, data were extracted from published figures using PlotDigitizer (https://plotdigitizer.com). This tool has been validated for accurate numerical extraction, with a Pearson correlation coefficient of 0.982 between true and extracted values[1].

*Machine learning classifiers*

Supervised learning classifiers were developed to categorize samples according to complement blockade. Observations were labelled as Inhibition = True when free C5 concentrations were ≤0.5 μg $ml^{-1}$, and Inhibition = False when >0.5 μg $ml^{-1}$. This threshold has been previously validated as an indicator of complete complement inhibition[2]. Algorithms tested included K-Nearest Neighbors (KNN), Logistic Regression (LR), Gradient Boosting (GB), Random Forest (RF), Support Vector Machines (SVM), Gaussian Naïve Bayes (GNB), and Extreme Gradient Boosting (XGBoost). Models were implemented in Python (RRID:SCR_008394, v3.9.12)[3] using scikit-learn (RRID:SCR_002577, v1.2.0)[4] and XGBoost (RRID:SCR_021361, v1.6.0)[5]. To estimate the concentration of eculizumab required for sustained inhibition, classifiers were queried with input values ranging from 10 to 1700 μg $ml^{-1}$, and the lowest concentration yielding ≥80% predicted probability of inhibition was recorded.

*Performance Measures*

Model predictive performance was evaluated using accuracy, precision, recall, and F1-score. Accuracy is defined as the proportion of correctly classified samples out of the total samples.

$$Accuracy = 1 - \frac{Number\ of\ misclassified\ samples}{Total\ number\ of\ samples}$$

Precision measures how well the model correctly identifies positive samples. In this case, a true positive is a positive sample that's correctly classified. A false positive happens when a negative sample is wrongly classified as positive. A true negative is a correctly classified negative sample, and a false negative is a positive sample incorrectly classified as negative.

$$Precision = \frac{True\ positives}{True\ positives + False\ positives}$$

The model's capability to identify positive samples from all possible positives is measured using recall, calculated as:

$$Recall = \frac{True\ positives}{True\ positives + False\ negatives}$$

The F1-score is the harmonic mean of precision and recall. The closer the F1-score is to 1, the better the precision and recall values. The F1-score is expressed as:

$$F1 - score = 2 \times \frac{Precision \times Recall}{Precision + Recall}$$

*Synthetic data generation*

To address the inherent scarcity of rare-disease datasets, we applied multiple synthetic data generation strategies as a conscious methodological innovation. Synthetic datasets were generated using Tabular Preset, Gaussian Copula, CTGAN, and TVAE methods from the Synthetic Data Vault (v0.18)[7], as well as the MOSTLY AI platform (https://mostly.ai[8]. Fidelity to real data was evaluated with the TableEvaluator library (v1.5.0) (https://baukebrenninkmeijer.github.io/table-evaluator/), which quantifies similarity across statistical distributions. In addition, synthetic datasets were assessed for utility by their ability to improve predictive performance when used for classifier training under controlled conditions of equal sample size and class distribution.

**Supplementary Table S1. Eculizumab-related data used for the model**

| Source | Disease | Eculizumab Concentration in steady state (μg/mL) | Free C5 concentration in steady state (μg/mL) |
|---|---|---|---|
| **C08-002A/B C08-003A/B NCT00844545 NCT00838513** | aHUS | 5.882 | 93.529 |
| | | 10.78 | 76.588 |
| | | 11.765 | 121.059 |
| | | 12.745 | 100.941 |
| | | 12.745 | 97.412 |
| | | 12.745 | 66 |
| | | 13.725 | 95.294 |
| | | 20.639 | 41.538 |
| | | 21.569 | 53.647 |
| | | 21.569 | 41.647 |
| | | 22.549 | 54 |
| | | 23.529 | 60 |
| | | 24.51 | 44.118 |
| | | 24.51 | 40.235 |
| | | 24.57 | 43.314 |
| | | 27.451 | 48.653 |
| | | 27.451 | 33.529 |
| | | 29.412 | 40.235 |
| | | 30.392 | 52.941 |
| | | 30.392 | 38.824 |
| | | 30.392 | 33.529 |
| | | 31.373 | 42.706 |
| | | 31.373 | 40.931 |
| | | 31.373 | 40.235 |
| | | 31.373 | 39.529 |
| | | 32.432 | 42.249 |
| | | 34.314 | 48.706 |
| | | 34.314 | 43.412 |
| | | 35.294 | 35.294 |
| | | 35.294 | 28.235 |
| | | 36.275 | 34.235 |
| | | 40.196 | 17.647 |
| | | 41.176 | 35.294 |
| | | 41.176 | 22.941 |
| | | 43.137 | 28.588 |
| | | 43.26 | 27.692 |
| | | 44.226 | 24.142 |
| | | 45.098 | 33.529 |
| | | 45.098 | 26.824 |
| | | 46.078 | 35.647 |

| | | |
|---|---|---|
| | 46.078 | 34.588 |
| | 46.078 | 28.941 |
| | 46.078 | 27.176 |
| | 46.078 | 23.294 |
| | 46.078 | 18 |
| | 46.192 | 28.402 |
| | 50 | 33.176 |
| | 50.123 | 29.822 |
| | 50.98 | 46.941 |
| | 50.98 | 24.706 |
| | 50.98 | 23.697 |
| | 51.106 | 29.822 |
| | 51.106 | 23.787 |
| | 51.123 | 27.692 |
| | 51.961 | 29.647 |
| | 52.941 | 28.235 |
| | 53.071 | 12.426 |
| | 53.922 | 12.706 |
| | 54.037 | 24.852 |
| | 54.054 | 26.627 |
| | 54.902 | 19.412 |
| | 55.037 | 11.361 |
| | 55.037 | 14.201 |
| | 56.863 | 31.059 |
| | 57.002 | 30.533 |
| | 59 | 31.059 |
| | 63.725 | 16.235 |
| | 64.706 | 27.176 |
| | 64.706 | 27.176 |
| | 64.706 | 21.882 |
| | 64.706 | 12.706 |
| | 64.865 | 14.201 |
| | 64.865 | 12.071 |
| | 65.686 | 23.297 |
| | 65.848 | 14.911 |
| | 66.83 | 14.201 |
| | 66.83 | 13.136 |
| | 66.83 | 12.071 |
| | 68.796 | 19.172 |
| | 69.608 | 13.412 |
| | 69.779 | 18.462 |
| | 70.588 | 18.7 |
| | 70.588 | 11.294 |
| | 70.762 | 20.947 |
| | 70.762 | 14.911 |

| | |
|---|---|
| 70.762 | 18.817 |
| 71.744 | 16.686 |
| 72.549 | 14.471 |
| 73.71 | 20.592 |
| 74.51 | 11.294 |
| 74.693 | 25.917 |
| 74.693 | 17.751 |
| 74.693 | 10.651 |
| 74.693 | 19.527 |
| 75.49 | 18.353 |
| 75.49 | 15.529 |
| 76.471 | 24 |
| 76.471 | 9.882 |
| 78.431 | 11.647 |
| 78.624 | 23.787 |
| 78.624 | 9.586 |
| 79 | 26.118 |
| 80.392 | 24 |
| 80.392 | 9.176 |
| 80.59 | 7.811 |
| 81.373 | 13.0559 |
| 81.572 | 13.846 |
| 83.333 | 14.824 |
| 83.538 | 22.367 |
| 83.538 | 16.686 |
| 84.314 | 23.294 |
| 84.314 | 16.941 |
| 84.521 | 13.846 |
| 86.486 | 7.811 |
| 88.235 | 11.294 |
| 89.435 | 15.621 |
| 90.418 | 15.266 |
| 91.4 | 7.811 |
| 95.332 | 6.746 |
| 96.314 | 5.68 |
| 96.314 | 8.521 |
| 96.314 | 13.136 |
| 99.02 | 8.471 |
| 100.246 | 9.231 |
| 101.229 | 8.166 |
| 102.2941 | 9.529 |
| 104.177 | 8.876 |
| 104.177 | 13.136 |
| 105.106 | 8.521 |
| 105.882 | 8.471 |

| | | |
|---|---|---|
| | 107.843 | 14.118 |
| | 108.824 | 15.88 |
| | 109.091 | 9.586 |
| | 109.091 | 8.166 |
| | 109.091 | 13.846 |
| | 110.074 | 7.811 |
| | 110.784 | 10.941 |
| | 112.039 | 19.172 |
| | 114.005 | 7.811 |
| | 115.972 | 17.751 |
| | 118.627 | 9.882 |
| | 118.919 | 6.036 |
| | 118.919 | 12.071 |
| | 122.85 | 13.136 |
| | 123.833 | 11.361 |
| | 124.816 | 7.101 |
| | 124.816 | 3.55 |
| | 124.816 | 4.615 |
| | 124.816 | 8.521 |
| | 125.799 | 4.97 |
| | 125.799 | 17.396 |
| | 127.764 | 6.746 |
| | 127.764 | 13.846 |
| | 128.431 | 5.294 |
| | 128.747 | 6.746 |
| | 129.412 | 7.765 |
| | 129.73 | 7.101 |
| | 129.73 | 6.746 |
| | 130.713 | 11.361 |
| | 130.716 | 5.68 |
| | 131.695 | 9.231 |
| | 131.695 | 20.237 |
| | 132.353 | 18 |
| | 132.678 | 5.68 |
| | 133.661 | 4.615 |
| | 133.661 | 5.325 |
| | 135.627 | 17.751 |
| | 137.592 | 12.426 |
| | 139 | 12.7 |
| | 139.216 | 11.294 |
| | 140.196 | 11.294 |
| | 141.523 | 4.615 |
| | 142.506 | 16.686 |
| | 143.137 | 9.882 |
| | 144.472 | 7.101 |

| | | |
|---|---|---|
| | 145.455 | 5.325 |
| | 147.059 | 14.118 |
| | 147.42 | 7.811 |
| | 148.039 | 9.176 |
| | 148.403 | 7.811 |
| | 149.02 | 8.118 |
| | 149.386 | 7.811 |
| | 149.386 | 12.781 |
| | 150 | 8.824 |
| | 150 | 8.118 |
| | 150.369 | 3.55 |
| | 150.369 | 8.166 |
| | 150.369 | 8.876 |
| | 150.369 | 14.201 |
| | 152.334 | 9.231 |
| | 153.317 | 6.036 |
| | 153.317 | 10.296 |
| | 154.283 | 8.521 |
| | 154.902 | 8.824 |
| | 159.214 | 6.036 |
| | 159.214 | 9.231 |
| | 160.784 | 7.059 |
| | 162.162 | 8.166 |
| | 164.128 | 8.521 |
| | 168.627 | 9.529 |
| | 170.025 | 9.941 |
| | 171.007 | 9.941 |
| | 174 | 9.529 |
| | 174.939 | 9.231 |
| | 176.904 | 9.586 |
| | 179.853 | 8.876 |
| | 179.853 | 6.036 |
| | 181 | 7.765 |
| | 181.818 | 8.521 |
| | 189.681 | 6.036 |
| | 193.612 | 7.101 |
| | 194.595 | 10.296 |
| | 200.491 | 4.26 |
| | 206.388 | 6.391 |
| | 206.863 | 7.059 |
| | 209.337 | 5.68 |
| | 210.319 | 10.296 |
| | 211.765 | 8.118 |
| | 213.268 | 6.391 |
| | 219.165 | 7.101 |

| | | | |
|---|---|---|---|
| | | 220.588 | 7.412 |
| | | 227 | 91.598 |
| | | 239.803 | 2.84 |
| | | 245.098 | 9.882 |
| | | 245.7 | 9.231 |
| | | 249.614 | 3.55 |
| | | 251.961 | 4.588 |
| | | 277.451 | 7.059 |
| | | 288.235 | 7.412 |
| | | 305.882 | 6 |
| | | 316 | 7.059 |
| | | 317 | 7 |
| | | 329 | 4.235 |
| **Ref.** [9] | gMG | 0.08722747 | 120 |
| | | 328.7 | 2.39 |
| | | 124.8 | 1 |
| | | 453.85 | 0.49 |
| | | 367.57 | 0.68 |
| | | 843.13 | 0.017 |
| | | 356.18 | 3.62 |
| | | 844.94 | 0.65 |
| | | 322.74 | 1.74 |
| | | 775.43 | 0.54 |
| | | 363.23 | 1.55 |
| | | 874.74 | 0.12 |
| | | 317.39 | 1.1 |
| | | 765.91 | 0.021 |
| **NCT01892345** | NMOSD | 4.06071654 | 297.275774 |
| | | 0 | 255.906383 |
| | | 0 | 252.148263 |
| | | 1.12111087 | 214.721892 |
| | | 2.29518761 | 208.496941 |
| | | 1.12993852 | 198.626613 |
| | | 0.3354505 | 192.739893 |
| | | 1.87146067 | 188.225638 |
| | | 2.97491625 | 185.01921 |
| | | 0 | 182.42761 |
| | | 3.16912443 | 178.060674 |
| | | 1.64194191 | 177.382709 |
| | | 0 | 172.169444 |
| | | 4.6256858 | 167.211731 |
| | | 8.03315663 | 161.733895 |
| | | 0 | 157.359443 |
| | | 1.30649141 | 154.047787 |
| | | 1.30649141 | 150.614368 |

| | | |
|---|---|---|
| | 1.28000847 | 147.843881 |
| | 0.40607165 | 143.830208 |
| | 0 | 140.444893 |
| | 0 | 137.190361 |
| | 17.4963917 | 134.510069 |
| | 0 | 133.584069 |
| | 0.67972864 | 130.555023 |
| | 0 | 127.449313 |
| | 0 | 124.713401 |
| | 0 | 121.573115 |
| | 0 | 117.577481 |
| | 0 | 114.515365 |
| | 1.74787364 | 108.53845 |
| | 0 | 106.205409 |
| | 0 | 104.163998 |
| | 0 | 101.864028 |
| | 0 | 100.260062 |
| | 0 | 98.6560961 |
| | 0 | 96.614685 |
| | 0 | 94.4274588 |
| | 0 | 92.6776779 |
| | 0 | 91.2195271 |
| | 0 | 89.3254344 |
| | 0 | 86.700763 |
| | 22.8812549 | 84.9765372 |
| | 0 | 80.7028025 |
| | 22.854772 | 76.4531198 |
| | 0 | 72.1312812 |
| | 0 | 69.0691645 |
| | 0 | 62.9569572 |
| | 16.3134873 | 63.2546004 |
| | 31.5058638 | 57.4896435 |
| | 58.3065929 | 0.12234477 |
| | 88.7178288 | 0.26515335 |
| | 109.736451 | 0.12384801 |
| | 131.258248 | 0.17195196 |
| | 149.902234 | 0.17044871 |
| | 179.757328 | 0.31810696 |
| | 204.580665 | 0.16593896 |
| | 247.809641 | 0.16443572 |
| | 265.791553 | 0.31025079 |
| | 283.773465 | 0.30874754 |
| | 343.209996 | 0.01260764 |
| | 374.309789 | 0.01260764 |
| | 413.58398 | 0.15842272 |

| | | | |
|---|---|---|---|
| | | 448.929869 | 0.26665659 |
| | | 471.661054 | 0.26665659 |
| | | 496.63446 | 0.37639372 |
| | | 523.92071 | 0.57782898 |
| | | 561.561787 | 0.57632573 |
| | | 596.748778 | 0.4290074 |
| | | 631.741562 | 0.4290074 |
| | | 672.207485 | 0.22606889 |
| | | 707.00606 | 0.37188397 |
| | | 742.193052 | 0.03515637 |
| | | 792.316418 | 0.18097145 |
| | | 816.82196 | 0.76423176 |
| | | 842.121989 | 0.26665659 |
| | | 864.058686 | 0.26665659 |
| | | 906.687382 | 0.2606436 |
| | | 929.886432 | 0.40645868 |
| | | 964.746801 | 0.20051367 |
| | | 1001.45215 | 0.45606587 |
| | | 1042.71256 | 0.3042378 |
| | | 1081.98675 | 0.15691947 |
| | | 1113.934 | 0.2636501 |
| | | 1142.27074 | 0.56730624 |
| | | 1174.63288 | 0.17379514 |
| | | 1215.42543 | 0.2636501 |
| | | 1244.23003 | 0.46508536 |
| | | 1289.25985 | 0.61090044 |
| | | 1329.34618 | 0.2200559 |
| | | 1361.96433 | 0.22189908 |
| | | 1395.35048 | 0.22189908 |
| | | 1426.25606 | 0.16928539 |
| | | 1464.40032 | 0.17078864 |
| | | 1507.35563 | 0.26665659 |
| | | 1547.01824 | 0.26665659 |
| **NCT02003144** | | 1586.48664 | 0.61725336 |
| | | 1606.18112 | 0.47444478 |
| | | 1650.54003 | 0.8833283 |
| | | 1692.36541 | 0.15425291 |
| | | 1717.27702 | 0.30157123 |
| | | 1755.64197 | 0.09829279 |
| **Ref**[10] | PNH | 0.0000 | 0.0520 |
| | | 216.0000 | 0.0520 |
| | | 108.0000 | 0.0460 |
| | | 162.0000 | 0.0440 |
| | | 310.0000 | 0.0410 |
| | | 216.0000 | 0.0460 |

| | | | |
|---|---|---|---|
| | | 229.0000 | 0.0350 |
| | | 229.0000 | 0.0440 |
| | | 229.0000 | 0.0180 |
| | | 472.0000 | 0.0520 |
| | | 229.0000 | 0.0460 |
| | | 229.0000 | 0.0460 |
| | | 243.0000 | 0.0590 |
| | | 243.0000 | 0.0490 |
| | | 445.0000 | 0.0660 |
| | | 216.0000 | 0.0490 |
| | | 216.0000 | 0.0590 |
| | | 229.0000 | 0.0790 |
| | | 243.0000 | 0.0440 |
| | | 367.0000 | 0.0520 |
| | | 489.0000 | 0.0520 |
| | | 353.0000 | 0.0460 |
| | | 285.0000 | 0.0440 |
| | | 557.0000 | 0.0410 |
| | | 367.0000 | 0.0460 |
| | | 285.0000 | 0.0350 |
| | | 272.0000 | 0.0440 |
| | | 272.0000 | 0.0180 |
| | | 272.0000 | 0.0520 |
| | | 503.0000 | 0.0460 |
| | | 272.0000 | 0.0460 |
| | | 272.0000 | 0.0590 |
| | | 244.0000 | 0.0490 |
| | | 272.0000 | 0.0660 |
| | | 517.0000 | 0.0490 |
| | | 244.0000 | 0.0590 |
| | | 244.0000 | 0.0790 |
| | | 299.0000 | 0.0440 |
| | | 285.0000 | 0.0660 |
| **Ref[11]** | | 0.0000 | 140.0000 |
| **Ref[12]** | | 0.0000 | 316.76 |

### Supplementary Table S2. TRIPOD-AI checklist.

| Section/Topic | | Checklist Item | Page |
|---|---|---|---|
| **Title and abstract** | | | |
| Title | 1 | Identify the study as developing and/or validating a multivariable prediction model, the target population, and the outcome to be predicted. | 1 |
| Abstract | 2 | Provide a summary of objectives, study design, setting, participants, sample size, predictors, outcome, statistical analysis, results, and conclusions. | 2 |
| **Introduction** | | | |
| Background and objectives | 3a | Explain the medical context (including whether diagnostic or prognostic) and rationale for developing or validating the multivariable prediction model, including references to existing models. | 3-4 |
| | 3b | Specify the objectives, including whether the study describes the development or validation of the model or both. | 3-4 |
| **Methods** | | | |
| Source of data | 4a | Describe the study design or source of data (e.g., randomized trial, cohort, or registry data), separately for the development and validation data sets, if applicable. | 7 |
| | 4b | Specify the key study dates, including start of accrual; end of accrual; and, if applicable, end of follow-up. | SI 3-11 |
| Participants | 5a | Specify key elements of the study setting (e.g., primary care, secondary care, general population) including number and location of centres. | 7 |
| | 5b | Describe eligibility criteria for participants. | 7 |
| | 5c | Give details of treatments received, if relevant. | 7 |
| Outcome | 6a | Clearly define the outcome that is predicted by the prediction model, including how and when assessed. | 7 |
| | 6b | Report any actions to blind assessment of the outcome to be predicted. | 7 |
| Predictors | 7a | Clearly define all predictors used in developing or validating the multivariable prediction model, including how and when they were measured. | SI 3-11 |
| | 7b | Report any actions to blind assessment of predictors for the outcome and other predictors. | SI |
| Sample size | 8 | Explain how the study size was arrived at. | 7 |
| Missing data | 9 | Describe how missing data were handled (e.g., complete-case analysis, single imputation, multiple imputation) with details of any imputation method. | NA |
| Statistical analysis methods | 10a | Describe how predictors were handled in the analyses. | SI 3-11 |
| | 10b | Specify type of model, all model-building procedures (including any predictor selection), and method for internal validation. | 7 |
| | 10d | Specify all measures used to assess model performance and, if relevant, to compare multiple models. | |
| Risk groups | 11 | Provide details on how risk groups were created, if done. | NA |
| **Results** | | | |
| Participants | 3a | Describe the flow of participants through the study, including the number of participants with and without the outcome and, if applicable, a summary of the follow-up time. A diagram may be helpful. | NA |
| | 3b | Describe the characteristics of the participants (basic demographics, clinical features, available predictors), including the number of participants with missing data for predictors and outcome. | SI 3-11 |
| Model development | 4a | Specify the number of participants and outcome events in each analysis. | 4-5 |
| | 4b | If done, report the unadjusted association between each candidate predictor and outcome. | NA |
| Model specification | 5a | Present the full prediction model to allow predictions for individuals (i.e., all regression coefficients, and model intercept or baseline survival at a given time point). | 4 |
| | 5b | Explain how to the use the prediction model. | 4 |
| Model performance | 16 | Report performance measures (with CIs) for the prediction model. | 4-5 |

| | | | |
|---|---|---|---|
| **Discussion** | | | |
| Limitations | 18 | Discuss any limitations of the study (such as nonrepresentative sample, few events per predictor, missing data). | 6 |
| Interpretation | 19b | Give an overall interpretation of the results, considering objectives, limitations, and results from similar studies, and other relevant evidence. | 5-6 |
| Implications | 20 | Discuss the potential clinical use of the model and implications for future research. | 5-6 |
| **Other information** | | | |
| Supplementary information | 21 | Provide information about the availability of supplementary resources, such as study protocol, Web calculator, and data sets. | 3-11 |
| Funding | 22 | Give the source of funding and the role of the funders for the present study. | 13 |

**Supplemental Table S3. Performance and predictions from machine learning models trained and tested using combinations of real and synthetic data.** Performance results are presented based on 10-fold cross-validation. Parameters corresponding to the best performing classifiers (KNN at *n* = 5000-10,000 data points and RF at *n* = 5000 data points) are shown in boldface.

| | **KNN** | **LR** | **RF** | **GNB** | **SVM** | **GB** | **XGB** |
|---|---|---|---|---|---|---|---|
| **Size of each training class, n = 303** | | | | | | | |
| ≥ 80% probability of minimum eculizumab level for complement inhibition | 187 | 317 | 162 | 437 | 234 | 162 | 162 |
| Mean accuracy | 0.89 | 0.87 | 0.87 | 0.73 | 0.90 | 0.89 | 0.90 |
| F1 score Complement inhibition = F | 0.89 | 0.87 | 0.86 | 0.78 | 0.90 | 0.89 | 0.90 |
| F1 score Complement inhibition = T | 0.90 | 0.86 | 0.87 | 0.64 | 0.91 | 0.89 | 0.91 |
| **Size of each training class, n = 500** | | | | | | | |
| ≥ 80% probability of minimum eculizumab level for complement inhibition | 161 | 406 | 162 | 529 | 243 | 162 | 173 |
| Mean accuracy | 0.91 | 0.86 | 0.90 | 0.74 | 0.90 | 0.90 | 0.90 |
| F1 score Complement inhibition = F | 0.90 | 0.87 | 0.89 | 0.79 | 0.89 | 0.90 | 0.90 |
| F1 score Complement inhibition = T | 0.91 | 0.85 | 0.90 | 0.66 | 0.90 | 0.91 | 0.90 |
| **Size of each training class, n = 1500** | | | | | | | |
| ≥ 80% probability of minimum eculizumab level for complement inhibition | 161 | 320 | 156 | 414 | 228 | 161 | 205 |
| Mean accuracy | 0.92 | 0.87 | 0.91 | 0.76 | 0.89 | 0.92 | 0.92 |
| F1 score Complement inhibition = F | 0.92 | 0.87 | 0.91 | 0.80 | 0.89 | 0.92 | 0.92 |
| F1 score Complement inhibition = T | 0.92 | 0.86 | 0.91 | 0.69 | 0.90 | 0.93 | 0.92 |
| **Size of each training class, n = 5000** | | | | | | | |
| ≥ 80% probability of minimum eculizumab level for complement inhibition | **162** | 310 | **152** | 395 | 223 | 108 | 162 |
| Mean accuracy | **0.94** | 0.88 | **0.94** | 0.76 | 0.91 | 0.93 | 0.93 |
| F1 score Complement inhibition = F | **0.94** | 0.89 | **0.94** | 0.80 | 0.91 | 0.93 | 0.93 |
| F1 score Complement inhibition = T | **0.94** | 0.88 | **0.94** | 0.70 | 0.91 | 0.94 | 0.93 |
| **Size of each training class, n = 10,000** | | | | | | | |

| | | | | | | | |
|---|---|---|---|---|---|---|---|
| ≥ 80% probability of minimum eculizumab level for complement inhibition | **162** | 319 | 160 | 401 | 223 | 162 | 162 |
| Mean accuracy | **0.94** | 0.88 | 0.93 | 0.76 | 0.90 | 0.93 | 0.93 |
| F1 score<br>Complement inhibition = F | **0.94** | 0.88 | 0.93 | 0.80 | 0.90 | 0.93 | 0.92 |
| F1 score<br>Complement inhibition = T | **0.94** | 0.87 | 0.94 | 0.70 | 0.91 | 0.93 | 0.93 |
